# LymphoML: An interpretable artificial intelligence-based method identifies morphologic features that correlate with lymphoma subtype

**DOI:** 10.1101/2023.03.14.23287143

**Authors:** Vivek Shankar, Xiaoli Yang, Vrishab Krishna, Brent T. Tan, Oscar Silva, Rebecca Rojansky, Andrew Y. Ng, Fabiola Valvert, Edward L. Briercheck, David M. Weinstock, Yasodha Natkunam, Sebastian Fernandez-Pol, Pranav Rajpurkar

## Abstract

Lymphomas vary in terms of clinical behavior, morphology, and response to therapies and thus accurate classification is essential for appropriate management of patients. In this study, using a set of 670 cases of lymphoma obtained from a center in Guatemala City, we propose an interpretable machine learning method, LymphoML, for lymphoma subtyping into eight diagnostic categories. LymphoML sequentially applies steps of (1) object segmentation to extract nuclei, cells, and cytoplasm from hematoxylin and eosin (H&E)-stained tissue microarray (TMA) cores, (2) feature extraction of morphological, textural, and architectural features, and (3) aggregation of per-object features to create patch-level feature vectors for lymphoma classification. LymphoML achieves a diagnostic accuracy of 64.3% (AUROC: 85.9%, specificity: 88.7%, sensitivity: 66.9%) among 8 lymphoma subtypes using only H&E-stained TMA core sections, at a level similar to experienced hematopathologists. We find that the best model’s set of nuclear and cytoplasmic morphological, textural, and architectural features are most discriminative for diffuse large B-cell lymphoma (F1: 78.7%), classic Hodgkin lymphoma (F1 score: 74.5%), and mantle cell lymphoma (F1: 71.0%). Nuclear shape features provide the highest diagnostic yield, with nuclear texture, cytoplasmic, and architectural features providing smaller gains in accuracy. Finally, combining information from the H&E-based model together with the results of a limited set of immunohistochemical (IHC) stains resulted in a similar diagnostic accuracy (accuracy: 85.3%, AUROC: 95.7%, sensitivity: 84.5%, specificity: 93.5%) as with a much larger set of IHC stains (accuracy: 86.1%, AUROC: 96.7%, specificity: 93.2%, sensitivity: 86.0%). Our work suggests a potential way to incorporate machine learning tools into clinical practice to reduce the number of expensive IHC stains while achieving a similar level of diagnostic accuracy.

## Introduction

Lymphomas are abnormal proliferations derived from lymphocytes (Jamil and Mukkamalla 2021). Over 50 different types of lymphoma have been identified and these vary in clinical behavior from indolent ones that can be managed with a “watch and wait” approach to aggressive types that require prompt administration of high intensity chemotherapy (Swerdlow et al. 2016). The process of precisely diagnosing lymphomas requires knowledge of the clinical history (e.g. site of involvement, history of solid organ transplant, etc.) and morphologic evaluation of hematoxylin and eosin (H&E)-stained tissue by a trained pathologist (Swerdlow et al. 2016). After evaluation of the H&E-stained slide and relevant clinical information, one or a few diagnoses are deemed most likely and this guides additional ancillary testing in the form of immunohistochemical (IHC) stains, flow cytometry and in some cases cytogenetic and molecular studies (Wang and Zu 2017; Sun, Medeiros, and Young 2016). Unlike some fields of pathology, in which a definitive diagnosis is frequently possible by using the H&E-stained tissue alone, for lymphoma diagnosis, IHC stains or flow cytometry are essential in the vast majority of cases. This is because identifying the cell of origin for lymphomas (B-cell, T-cell, and NK cells) is essential for definitive diagnosis and treatment (Nowakowski et al. 2019), but this cannot be reliably determined based on the H&E-stained section alone. In contrast to the H&E-stained section, which is inexpensive and widely available, IHC stains and flow cytometry require costly equipment, expensive reagents, and trained personnel. In general, more experienced pathologists tend to require fewer ancillary studies, and thus greater experience may lead to more efficient resource utilization. However, worldwide, the shortage of pathologists is so great that modest improvements in the efficiency of pathologists is unlikely to make a significant impact in reducing the costs of lymphoma diagnosis (Metter et al. 2019; Eniu et al. 2017). Thus, cost-effective strategies that can help general pathologists reduce the number of ancillary studies may help to reduce the cost of lymphoma diagnosis.

Though this represents an oversimplification, pathology resources available in low-middle income countries (LMICs) may be divided into three general levels (Valvert et al. 2021; Fleming et al. 2017). At the lowest level, there may not even be resources for making H&E-stained sections. In other countries, there may be resources to generate H&E-stained slides but pathologists are limited in number and subspecialty expertise is lacking. Another level may be defined by the availability of more resource-intensive assays such as immunohistochemistry flow cytometry. Conceptually, AI tools may be useful in several scenarios in LMICs by 1) screening specimens to reduce the number of slides that require pathologist review, 2) allowing diagnoses to be made using H&E-stained sections alone, or 3) allowing general pathologists to maximize the diagnostic yield from the H&E to minimize the number of IHC-stained sections necessary to make an accurate diagnosis.

Machine learning tools applied to diagnostic pathology have shown great promise in analyzing H&E-stained images. Though the equipment necessary for scanning slides and computational algorithms are not yet in widespread use, hardware and software improve rapidly and thus machine learning tools hold promise for improving the accuracy and efficiency of diagnostic pathology. For the task of lymphoma diagnosis, machine learning studies have achieved high accuracies ranging from 94-100% when classifying between a small number of lymphoma subtypes (ranging from 2-4 diagnostic categories: diffuse large B-cell lymphoma (DLBCL), non-DLBCL (Li et al. 2020); DLBCL, Burkitt lymphoma (BL) (Mohlman et al. 2020); chronic lymphocytic leukemia (CLL), DLBCL, control lymph nodes (Steinbuss et al. 2021); DLBCL, follicular lymphoma (FL), reactive lymphoid hyperplasia (Miyoshi et al. 2020); CLL, FL, mantle cell lymphoma (MCL) (Janowczyk and Madabhushi 2016; Brancati et al. undefined 2019; Zhang et al. 2020); benign, DLBCL, BL and small lymphocytic lymphoma (Achi et al. 2019)). However, classifying between the aforementioned small number of lymphoma subtypes does not reflect the full scope of complexity encompassed in the workflows of pathologists. Tools that can accurately distinguish among a larger number of diagnostic categories may provide greater clinical value for diagnostic pathologists in real-world settings.

In this work, we introduce LymphoML, an interpretable machine learning approach for lymphoma subtyping into eight diagnostic categories. LymphoML sequentially applies steps of (1) object segmentation to extract nuclei, cells, and cytoplasm from H&E-stained TMA cores, (2) feature extraction of morphological, textural, and architectural features, and (3) aggregation of per-object features to create patch-level feature vectors for lymphoma classification. We apply LymphoML to a dataset of 670 lymphoma cases from Guatemala containing eight diagnostic categories. When using only the H&E-stained TMA core, we find that LymphoML outperforms deep-learning approaches, and demonstrates non-inferiority to an experienced hematopathologist and a general pathologist. We further evaluate the boost in model performance when combining the best model produced by LymphoML with a limited number of pathologist-interpreted immunostains. This latter assessment suggests a potential way to incorporate machine learning tools into the practice of pathology to reduce the number of expensive IHC stains while achieving a similar level of diagnostic accuracy.

## Materials and Methods

### Dataset

This study was approved by the institutional review board of Stanford University and La Liga Nacional Contra El Cáncer. The cases used for this study were selected and tissue microarrays (TMAs) were constructed as previously published (Valvert et al. 2021). The authors in (Valvert et al. 2021) retrospectively reviewed medical records to identify all formalin-fixed, paraffin-embedded (FFPE) biopsy specimens obtained at Instituto de Cancerologia y Hospital Dr. Bernardo Del Valle (INCAN) because of clinical suspicion of lymphoma between 2006 and 2018. One-half of each FFPE block was shipped to Stanford University for H&E whole-slide image generation. Two expert hematopathologists reviewed the slides, selected regions of interest (ROIs), and included two cores from each sample for tissue microarray (TMA) construction. The H&E-stained TMAs were scanned at 40x magnification (0.25 µm per pixel) on an Aperio AT2 scanner (Leica Biosystems, Nussloch, Germany) in ScanScope Virtual Slide (SVS) format. As described previously (Valvert et al. 2021), diagnoses were established based on the World Health Organization (WHO) classification (Swerdlow et al. 2016) and then binned into 8 categories: aggressive B-cell (Agg BCL), diffuse large B-cell (DLBCL), follicular (FL), classic Hodgkin (CHL), mantle cell (MCL), marginal zone (MZL), natural killer T-cell (NKTCL), or mature T-cell lymphoma (TCL). All of the TMA blocks (seven total) were also stained for 46-different markers by immunohistochemical (IHC) stains as described previously (Valvert et al. 2021). Each IHC stained TMA was assessed by a hematopathologist to determine if the lymphoma cells were positive or negative for the marker. The complete list of cases with the associated IHC results as interpreted by a hematopathologist is provided in Supplementary Table 2. The distribution of cases in each lymphoma subtype is provided in Supplementary Table 3. Summarized clinical data for the patients with suspected lymphoma and individual patient characteristics can be found in a previously published study’s Supplementary Tables 4 and 5 <u>(Valvert et al. 2021)</u>.

### Patch Extraction

The H&E-stained tissue cores were indicated by hematopathologists using Qupath (Bankhead et al. 2017). From each tissue core, we extracted a fixed number of patches at 40x magnification. Non-overlapping patches were extracted from inside each tissue core, starting from the top-left and proceeding until the bottom-right corner. We omitted patches that were mostly white and contained little tissue. Specifically, background was defined as pixels with saturation value less than 0.05 in HSV space, and we excluded patches where more than 95% of the pixels were background.

### Nuclei and Cell Segmentation

We considered two different deep-learning based nuclear segmentation models (HoVer-Net (Graham et al. 2019) and StarDist (Weigert et al. 2020)) to segment every nucleus inside each of the H&E-stained TMA cores. HoVer-Net uses a neural network based on a pre-trained ResNet-50 architecture to extract image features. StarDist is powered by a pre-trained deep-learning CNN that predicts a suitable shape representation (star-convex polygon) for each cell nucleus. We first normalized the input image pixel intensities to the range 0.0 to 1.0 using percentiles of 1 and 99 to clip the bottom and top 1% of pixel values to 0.0 or 1.0. Then, we ran StarDist segmentation, which operated independently on each TMA core and produced an output image segmenting all individual cell nuclei in the core.

We selected StarDist as the nuclei segmentation algorithm for all our cases. We measured the agreement of HoVer-Net’s nuclei segmentations with StarDist’s nuclei segmentations by computing the mean Intersection over Union (mIOU) over all segmented patches. We obtained a mIOU of 0.762, which is a high degree of agreement between HoVer-Net and StarDist segmentations. Additionally, we found that the best-performing H&E-only models utilizing features extracted from StarDist achieved marginally higher top-1 accuracy (64.3%) than the best-performing models using features extracted from HoVer-Net (61.5%). As both algorithms are widely used and in close agreement, we chose StarDist because it resulted in higher performance in downstream applications, though the improvement in performance was marginal.

### Feature Extraction

We used the per-nucleus binary segmentation masks output by StarDist to compute several geometric features for each cell nucleus using methods similar to those by (Vrabac et al. 2021). Using manually extracted features allowed our models to produce interpretable results, and facilitated identification of the features that were most important in driving the classification using methods such as SHAP (SHapley Additive exPlanations, described below) (Lundberg and Lee 2017). We calculated features such as minimum/maximum Feret diameters, convex hull area of the segmented nucleus, and a number of other derived geometric features including measures of circularity, elongation, and convexity. To obtain a richer feature set, we used CellProfiler (Carpenter et al. 2006), an open-source tool for analyzing biological images, to extract quantitative features of the morphology, color intensity, and texture of segmented nuclei and cells. We constructed an image analysis pipeline in CellProfiler consisting of modules to process the H&E cores, identify nuclear and cell boundaries, and measure features of the identified objects. First, a color deconvolution was performed on each image tile to create a set of separate hematoxylin and eosin stained images in grayscale. Next, we ran StarDist on the hematoxylin image to produce a binary mask segmenting the nuclei. We used the binary segmentation mask to identify nuclear objects; minimum and maximum diameters were configured for the sizes of objects to be identified. The primary objects (which in our study are nuclei) were subsequently used as a reference to identify secondary objects such as the cells and cytoplasm. Finally, we extracted size, shape (e.g. bounding box area, minor axis length (MAL)), color intensity, and texture (e.g. mean intensity, integrated intensity) features from the detected cells and nuclei. We specified options to calculate Zernike shape features using the first 10 Zernike polynomials (order 0 to order 9) and calculated advanced features (statistics for object moments and inertia tensors). The first 10 Zernike features were the default in CellProfiler and while there is no limit on the number of features, higher order polynomials carry less and less information. The full list of features extracted by CellProfiler is provided in Supplementary Table 4. Definitions for each feature are provided in the CellProfiler Measurement module (Stirling et al. 2021).

To obtain a single feature vector for each patient, each of the features extracted by CellProfiler was aggregated across all cells in a patch by their mean, standard deviation, skew, kurtosis, and percentiles, yielding a total of 1605 features for each patient.

### Spatial Relationship Features

To assess the spatial relationships between nuclei, we considered architectural features from two sources: 1) CPArch: features that contain architectural information provided by CellProfiler (e.g. BoundingBoxMaximum, Center, etc), and 2) CT: statistical spatial features representing clustering tendency (CT) characterized from the Ripley’s K function. When computing CT, we followed the steps described in (Subramanian et al. 2018). Specifically, we used centroid coordinates (in pixels) to define cell locations in each patch and computed values of the self-K function at different radii. The optimal radii range was determined by cross-validation. The resulting vector consisting of self-K function values at each radius was used as the patch’s CT feature. When performing lymphoma subtype predictions, we concatenated CPArch features and CT vectors directly to the rest of the features.

### Models

We used LightGBM (Ke et al. 2017), a fast and efficient tree-based machine learning algorithm that employs a gradient boosting framework, for all experiments. We handled class-imbalance by preserving the label distribution when splitting the dataset and made sure patches from the same patient were in the same data split. To correct the bias induced by class-imbalance during model training, we used focal loss (Lin et al. 2017) as the loss function, and turned on ‘balanced’ mode in LightGBM to automatically adjust weights inversely proportional to class frequencies in the input data. We used 5-fold cross-validation for all experiments.

To select the best-performing feature-based model, we performed patch-resolution experiments. We considered cases when each core was divided into a perfect square number of patches (e.g. 1, 4, 9, …, 100 patches). Using this patch extraction method, we divided the width and height of each TMA core into a fixed number of segments to produce a grid of equally-sized patches. Additionally, since TMA cores come in varying sizes, this method preserves the same label distribution in the patch-level dataset as in the original core-level dataset as we simply scale up the number of examples by a constant. We compared models fitted using features aggregated from patches of different sizes, selected the best model based on the 5-fold cross-validation accuracy, and reported the model’s test performance by its accuracy and weighted F1 scores, both overall and per-class.

### Feature Importances

We used the SHapley Additive exPlanation (SHAP) method to quantify the impact of each feature on the trained model (Lundberg and Lee 2017). The SHAP method explains prediction by allocating credit among the input features; feature credit is calculated using Shapley Values as the change in the expected value of the model’s predicted score for a label when a feature is present versus absent. We also grouped related morphological features into different categories and ran the SHAP method on the resulting feature groups. We summed the raw SHAP values within each group to get an estimate of the group’s importance as a whole.

### Deep Learning

We prepared image data by dividing cores into patches of 224x224 pixels with 50% overlap (a common form of data augmentation) and filtered these images as described in “Patch Extraction” with white patches discarded. Patch pixels were normalized and standardized to have mean 0 and variance 1. On these preprocessed patches, we implemented a set of deep-learning approaches to lymphoma classification. We fine-tuned two open-source models pretrained on H&E patches – a ResNet-50 self-supervised on a number of tasks and cancers with H&E and IHC stained slides (He et al. 2016) and a specialized TripletNet architecture pre-trained on CAMELYON16 (a dataset consisting of breast cancer H&E WSIs) (Srinidhi et al. 2022). We trained both models, adding a linear layer as the final classification layer and allowed weights in all layers to update during fine-tuning. Focal loss was used to handle class-imbalance with normalized weights generated from label proportions on the training set and a gamma parameter of 2.0. The model was updated by an Adam Optimizer (Kingma and Ba 2014) with a learning rate of 0.001 and a batch size of 128 for 100 epochs. We split our dataset at a core-level into training, validation, and test splits to ensure that all extracted image patches from the same patient are in the same data split with 70% of the total tissue microarray (TMA) cores for training, 10% for validation to tune model hyperparameters, and 20% for testing.

### Environment

TMA cores were processed in a CellProfiler (version 4.2.1) pipeline for object detection and feature measurement. Using Squidpy (version 1.1.2) (Palla et al. 2022), we ran a pre-trained StarDist model checkpoint for brightfield H&E images on each TMA core to generate a binary nuclei segmentation mask. We computed CT features using Ripley’s K function defined in spatstat (spatstat: version 1.63_3, spatstat.data: version 1.4_3) in R (version 4.1.2). Pre-trained TripletNet and ResNet deep-learning models were fine-tuned on four NVIDIA GeForce GTX 1070 Ti GPUs using Pytorch Lightning (version 1.4.9). Gradient boosting models were trained using lightgbm (version 3.3.1). Plots were generated in Python and R using matplotlib (version 3.4.3), seaborn (version 0.11.2), plotly (version 5.6.0), ggplot2 (version 4.1.0), and ComplexHeatmap (version 4.1.1). Additionally, the following Python libraries were used for analysis and data handling: h5py (3.4.0), numpy (1.21.2), opencv (3.4.2), openslide-python (1.1.2), pandas (1.3.3), pillow (8.3.2), scikit-learn (0.24.2), scipy (1.6.2), and shap (0.39.0). The 95% CIs for accuracy and F1 scores were computed using non-parametric bootstrapping from 1,000 bootstrap samples.

### Evaluation

We assessed the performance of LymphoML (index test) in predicting the ground-truth World Health Organization (WHO) diagnosis (reference standard) for each case. The index test results were not available to assessors of the reference standard. The assessors of the reference standard reviewed H&E slides generated from whole sections and IHC results to classify each specimen according to the WHO classification (Valvert et al 2021). We additionally compared the performance of LymphoML to those of human-benchmark pathologists. The H&E-stained TMA cores or WSIs were each reviewed by hematopathologists who were blinded to all other patient information and clinical data including the results of immunohistochemical stains and the final histopathologic diagnosis (reference standard results). Other than the H&E-stained tissue, no additional clinical information or reference standard results (i.e., pathologist-interpreted immunostain markers, results of histopathology) were provided to the human-benchmark pathologist or to LymphoML. There were no indeterminate index test or reference standard results.

We analyzed overall model performance by top-1 accuracy, F1 score (harmonic mean of precision and recall scores), AUROC, sensitivity, and specificity. We calculated metrics for each label individually and found their weighted average across all labels. We weight by the support, or the number of true instances for each label, to account for class imbalance. Let 𝑇𝑃_𝑖_, 𝐹𝑃_𝑖_ , 𝑇𝑁_𝑖_ and 𝐹𝑁_𝑖_ represent the number of true positives, false positives, true negatives, and false negatives for class _𝑖_. Then, we compute the precision, recall, and F1 scores of class _𝑖_ using:

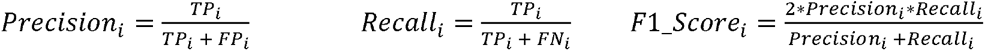

Let 𝑘 represent the total number of labels, 𝑛 represent the total number of examples, and |𝑦_𝑖_| represent the number of examples with label _𝑖_. We compute the weighted F1 score as follows.

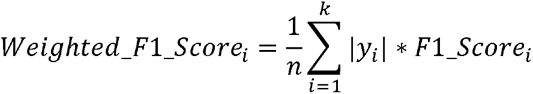

We followed a similar procedure to calculate weighted sensitivity, specificity, and AUROC metrics. We computed 95% CIs for all metrics using non-parametric bootstrapping from 1,000 bootstrap samples to account for uncertainties in our test results. These metrics are summarized in Supplementary Table 7 for all models using features extracted from H&E-stained tissue only. We performed statistical tests checking for any significant differences and equivalence (TOST) relationships by comparing the top-1 accuracy of the Best H&E Model with two pathologists on H&E-stained TMA cores and two pathologists on H&E-stained WSIs. Specifically, the two-tailed paired t-test between the Best H&E Model and Pathologist 1, for example, checks whether the mean difference for a particular metric (e.g. top-1 accuracy) between the two methods is zero. The significance level is 5%. Let 𝜇*_Best H&F Model_* represent the mean of a metric (e.g. mean top-1 accuracy) for the Best H&E Model and 𝜇*_pathologist_* represent the mean of a metric for the Pathologist. TOST consists of two one-tailed paired t-tests with the following null hypothesis:

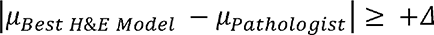

which is equivalent to two separate null hypotheses

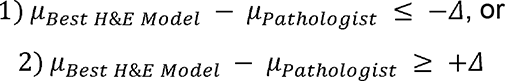

A TOST result that rejects both inequalities indicates that the mean difference between Best H&E Model and Pathologist metrics falls within the equivalence interval (−𝛥+𝛥). When only 1) is rejected, we conclude that the Best H&E Model is non-inferior to the Pathologist. The maximum clinically acceptable difference (𝛥) is designed by domain knowledge and was chosen to be 5%. Both the test for significant difference and TOST were performed by computing paired metric differences between two methods and deriving the 95%- (test for significant difference) and 90%-CIs (TOST) using 1000 bootstrap samples. Note that comparing the lower limit of the 90%-CI with −𝛥 is equivalent to performing a one-tailed test with 5% significance level. By comparing the lower limit of the 90%-CI with −𝛥 and the upper limit with +𝛥, we test null hypotheses 1) and 2), respectively. Test results and confidence intervals are summarized in Supplementary Table 12.

In addition to model-pathologist comparisons, we also used these statistical tests to perform model-model comparisons to evaluate the usefulness of specific sets of features. First, we compared models using nuclear morphological features only to models using additional intensity/textural features, architectural features, and cytoplasmic features. Next, we compared the performance of a baseline model using 46 different immunostains (and no H&E) to the Best H&E Model augmented with six hand-picked immunostains. This study was conducted in compliance with STARD guidelines (see Supplementary Table 1).

## Results

### Section 1: Interpretable Models for Lymphoma Diagnosis leveraging nuclear morphology and architecture

#### Section 1.1 Nuclear Morphology

We first tested whether nuclear shape features had higher diagnostic yield than nuclear texture or cytoplasmic features for accurately classifying lymphoma subtypes. The model using only nuclear features achieved 59.7% ([50.4%, 68.2%]) top-1 test accuracy, while models using nuclear texture or cytoplasmic features alone achieved slightly lower accuracy. Adding nuclear intensity or cytoplasmic features to the nuclear shape features marginally improved performance by another 1-2%. We analyzed model performance by lymphoma subtype using per-class F1 (harmonic mean of precision (specificity) and recall (sensitivity)) scores. We observed that nuclear features were most discriminative for diffuse large B-cell lymphoma (DLBCL) (F1 score: 76.2%), classic Hodgkin lymphoma (CHL) (F1 score: 65.3%), and mantle cell lymphoma (MCL) (F1 score: 51.6%) and were largely unable to distinguish other lymphoma subtypes (F1 scores less than 50.0%). To extract morphologic features from the H&E-stained images, we used the data processing, feature extraction pipeline, and modeling workflow described in the Materials and Methods section (Figure 1).

**Figure 1.**
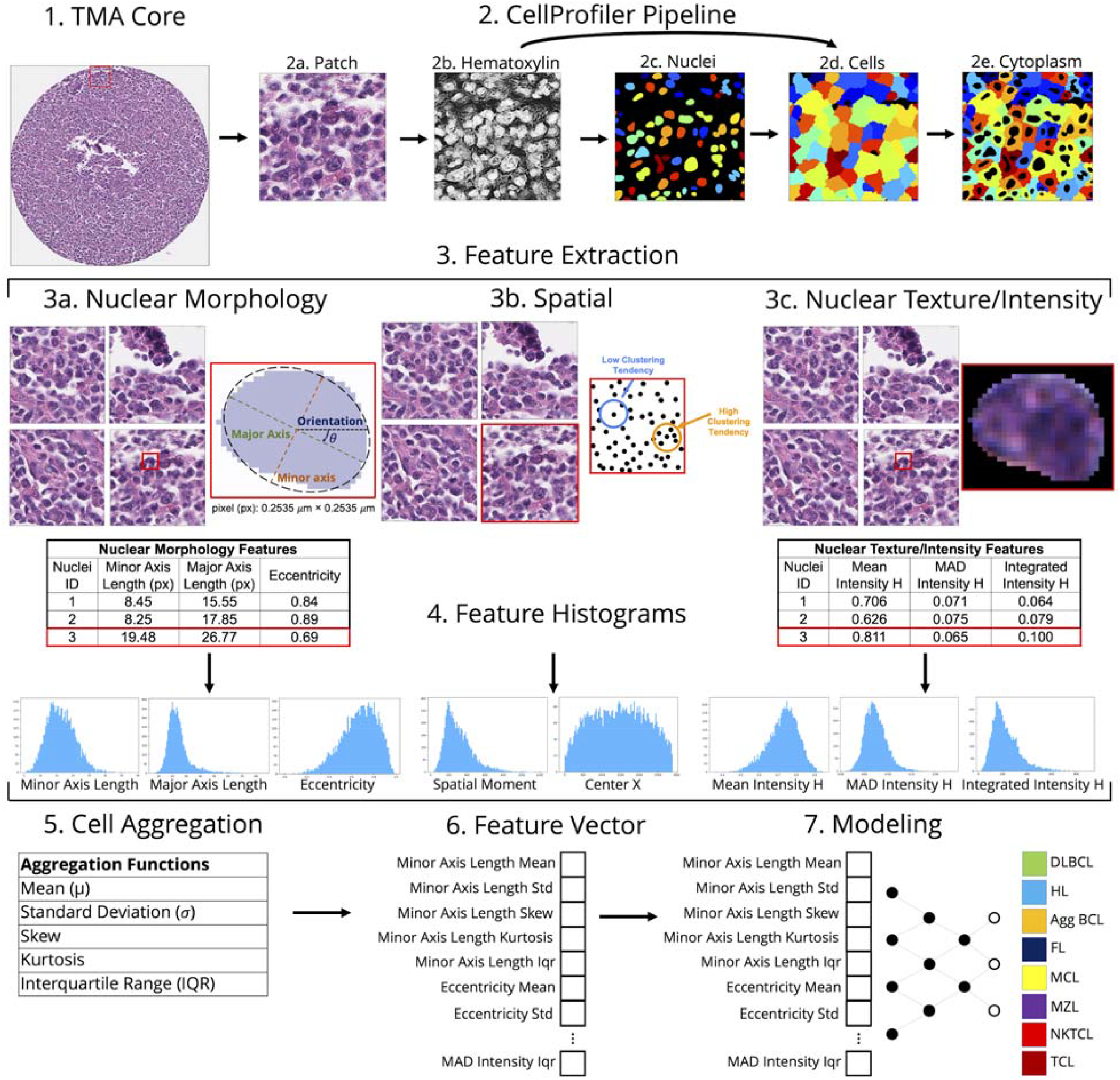
(Method Overview): (1) **Patch Extraction:** Patches of a fixed size are extracted from each TMA core (2) **CellProfiler Pipeline:** (a) Using CellProfiler, each patch was converted from the H&E-stained image into separate Hematoxylin (H) and Eosin (E) images. (b) H image (the E image was not required for the remainder of the pipeline). (c) We applied the StarDist algorithm on the H image to produce a nuclei segmentation mask. (d) Using the H image and the nuclei segmentation mask together, we used CellProfiler to identify the cell boundaries. We identified the cell boundary by using the nuclei as a “seed” region, then growing outwards until stopped by the H image threshold or by a neighboring nucleus. The Propagation method was used to delineate the boundary between neighboring cells. (e) We identified the cytoplasm by “subtracting” the nuclei objects from the cell objects. (3) **Feature Extraction/Measurement**: for each identified nucleus, cell, and cytoplasm, the following groups of features were extracted and measured: (a) nuclear morphology features, (b) spatial/architectural features (c) texture/intensity features. (4) **Feature Histograms:** Histograms for sample features in each feature group are displayed. The histograms display the distribution of a particular measured feature for all objects present in the entire patch. (5) **Cell Aggregation:** For each measurement obtained, the mean, standard deviation, skew, kurtosis, and interquartile range (IQR) were calculated for the entire population of objects present in a patch. We used these statistics to capture the distribution of each feature across the patch. (6) **Feature Vector:** The aggregated features were packed into a feature vector. (7) **Modeling:** Using the feature vector as input, models were trained to predict the most likely lymphoma subtype for the patch. The plurality vote across the patch-level model predictions was used to obtain the final core-level prediction.

**Figure 2.**
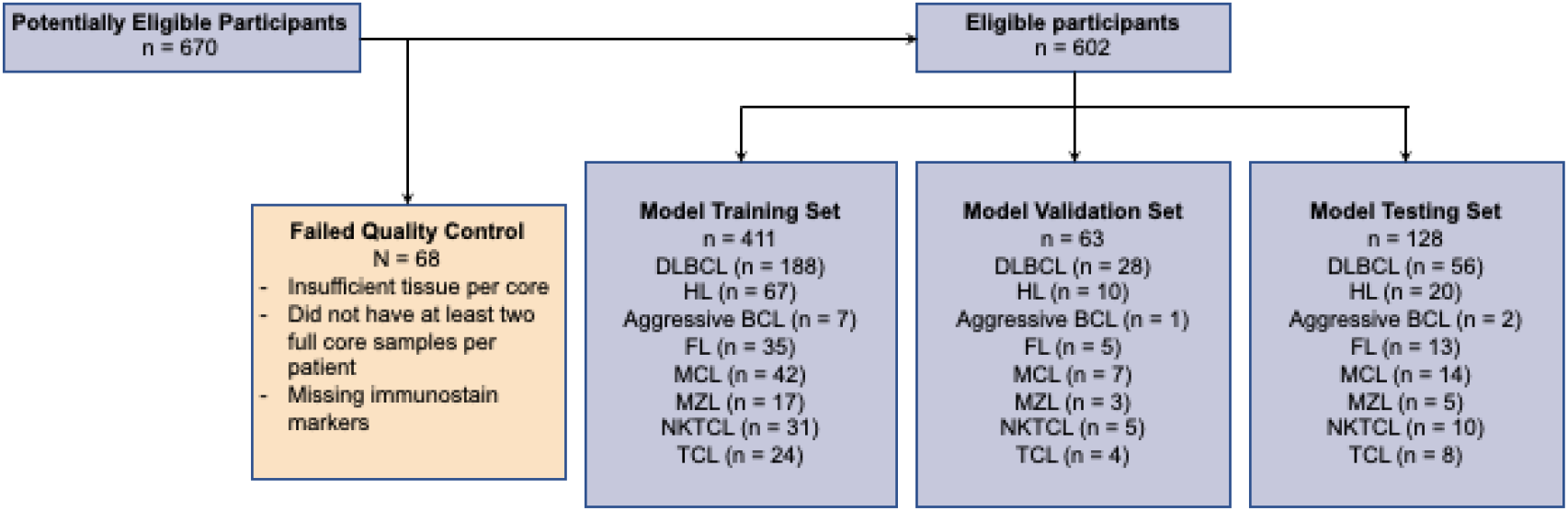
(Schema of samples from INCAN): Of 670 FFPE biopsy specimens, 68 failed quality control (e.g. did not have sufficient tissue per core, at least two full samples per patient, were missing ground-truth diagnoses, missing immunohistochemical stains, etc.) and were excluded from the dataset used to train and evaluate the model. The remaining 602 samples were split at a core-level into training, validation, and test splits to ensure that all extracted image patches from the same patient are in the same data split with 70% of the total tissue microarray (TMA) cores for training, 10% for validation to tune model hyperparameters, and 20% for testing. Stratified sampling was used to proportionally represent the eight diagnostic categories in each of the training, validation, and test sets.

We utilized the SHAP method to investigate whether area shape features played the biggest roles in classifying lymphoma subtypes among all nuclear features. We reported the resulting Shapley values for the top 20 nuclear features on individual predictions. We also trained and evaluated a parsimonious model using only the top eight most impactful features for each class as determined by SHAP; the resulting model achieved a top-1 test accuracy of 61.2% ([53.5%, 69.0%]) using just 10% of all features. The majority of the top 20 nuclear features identified by SHAP were nuclear area shape features such as mean radius, minor axis length, maximum feret diameter, solidity, orientation, maximum radius length, and nuclei area. In particular, MinorAxisLength is most helpful in diagnosing MCL and FL, and MeanRadius is most helpful in diagnosing CHL. Next, we grouped related morphological features into different categories and ran the SHAP method on the resulting feature groups. The nuclear size feature group had the largest mean absolute SHAP value, suggesting that of all nuclear features, size features were the most helpful for classifying DLBCL, CHL, and MCL (Figure 4).

We selected a few of the top features and analyzed their ability to distinguish patients with specific lymphoma subtypes. For example, the MinorAxisLength parameter was significantly different between cases of DLBCL and MCL. The minor axis length (MAL) is the length (in pixels) of the ellipse that has the same normalized second central moments as the nucleus region (Stirling et al. 2021). Heatmaps of the MAL for representative cases of DLBCL, CHL, and MCL (see Figure 3a) showed that DLBCL generally has cells with a larger minor axis length, which is consistent with the World Health Organization Classification system definition of DLBCL as a B-cell lymphoma with large cells (Swerdlow et al. 2016). The patch with the highest mean MAL taken from the DLBCL case had a higher mean MAL than the patch with the highest mean MAL taken from the CHL and MCL (see Figure 3b(iv)). Similarly, the patch with the lowest mean MAL taken from the DLBCL case had a higher mean MAL than the patch with the lowest mean MAL from the CHL and MCL cases (see Figure 3b(iv)). Assessment of the MAL for all patches in the core also showed that the MAL is higher for the DLBCL case than the MAL for all patches in the core for CHL and MCL (see Figure 3b(v)).

**Figure 3a.**
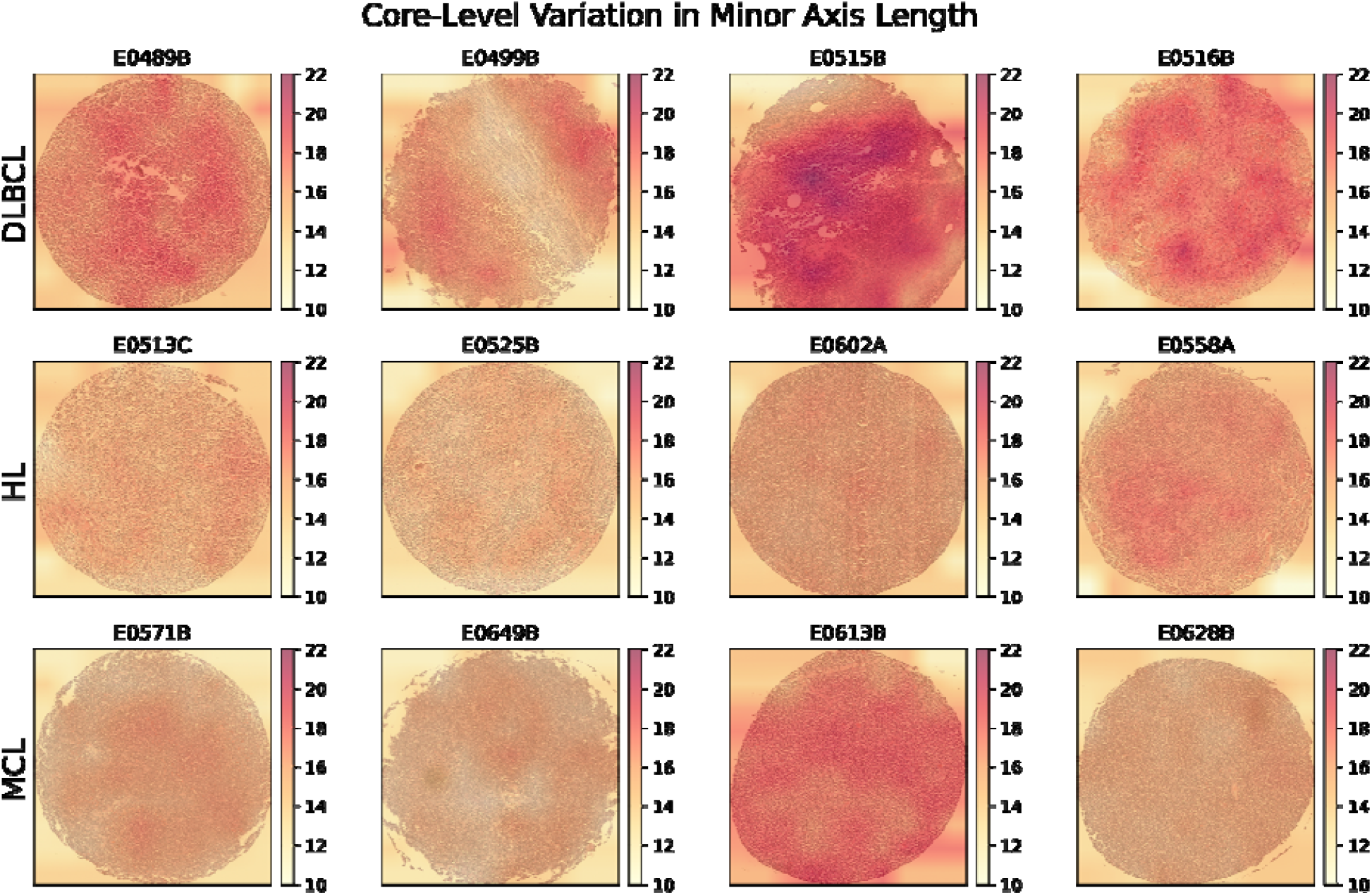
(Core-Level Variation in Minor Axis Length): The minor axis length (MAL) is the length (in pixels) of the ellipse that has the same normalized second central moments as the nucleus region. The MAL for each patch is shown as a heatmap superimposed on selected TMA cores of four cases each of diffuse large B-cell lymphoma (DLBCL), classic Hodgkin lymphoma (CHL), and mantle cell lymphoma (MCL). The nuclei of DLBCL cells generally have higher MAL.

**Figure 3b.**
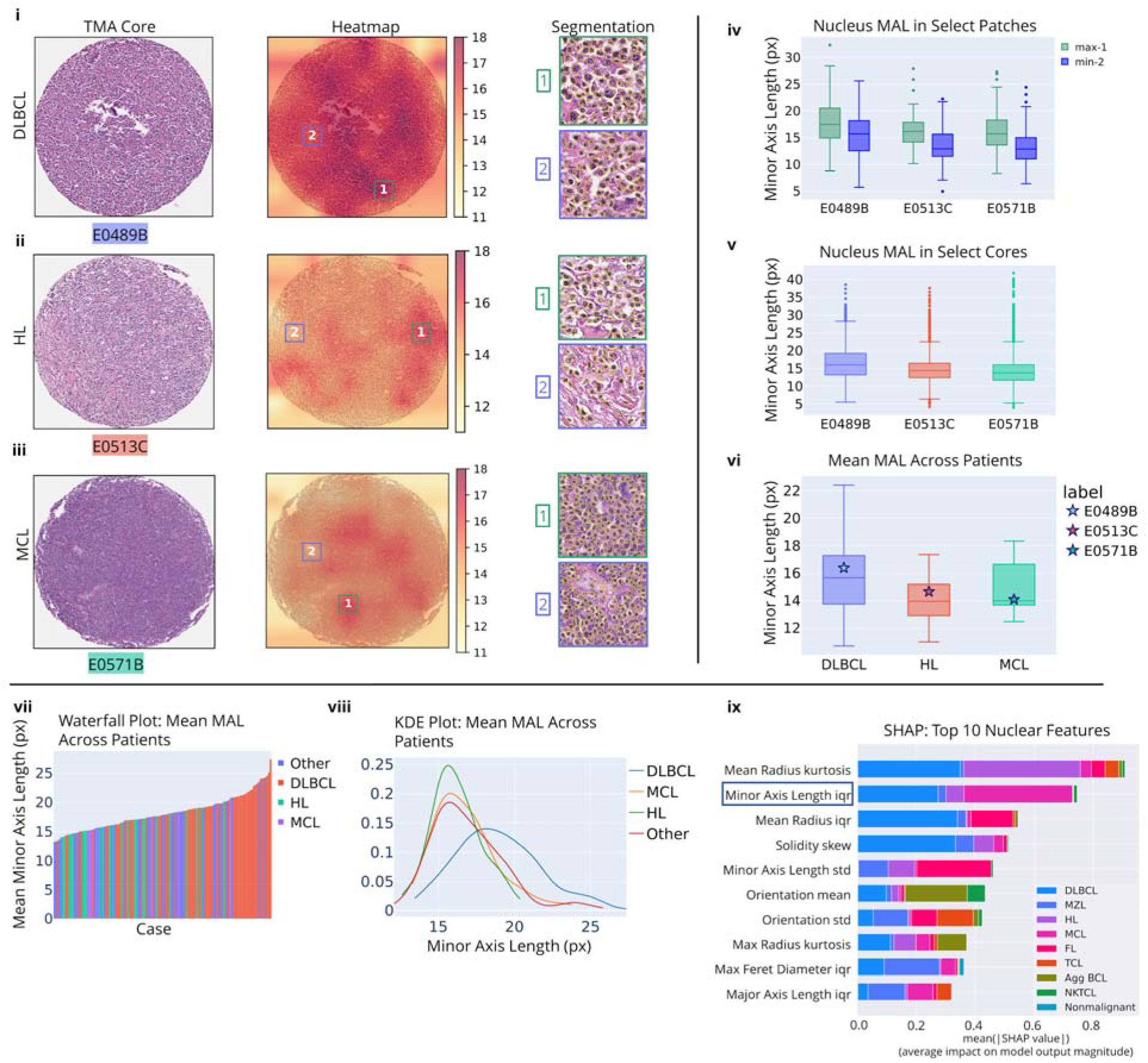
(Minor Axis Length Feature Analysis): Comparison of Minor Axis Length (MAL) across lymphoma subtypes. The minor axis length is the length (in pixels) of the ellipse that has the same normalized second central moments as the identified nucleus region. For selected cases from (i) DLBCL, (ii) CHL, and (iii) MCL, we plot a heatmap superimposed on the tissue microarray (TMA) cores showing the variability in nuclei minor axis length across the core. We also mark the patches with the largest (1) and smallest (2) mean MAL. For each patch, we display the segmented nuclei using StarDist nuclei segmentation. The maximum and minimum patches show visually the variability in nuclei size in a specific core. In (iv), we show the distribution of nuclei MAL in the maximum (1) and minimum (2) **patches** for each case. In (v), we show the distribution of nuclei MAL in the overall **cores**. In (vi), we show the distribution of mean MAL across patients in the entire **TMA**. DLBCL cases generally have the largest nuclei and greater variability in nuclei size than either CHL or MCL cases. In (vii), we display a waterfall plot of the mean minor axis length across patients. For each case/patient, we plot the mean of the top five patches with the highest minor axis length. DLBCL cases generally have the largest mean minor axis lengths of all lymphoma subtypes. In (viii), we show a Kernel density estimation plot. The distribution of minor axis length is clearly further to the right for DLBCL than for CHL and MCL. Finally, in (ix), we show a feature importance analysis plot by SHapley Additive exPlanation (SHAP) values. The top 20 nuclear features by percentage importance using the SHAP method are shown: minor axis length interquartile range (IQR) is the second-most important feature identified by SHAP. The majority of top 20 nuclear features are area-shape features.

**Figure 4.**
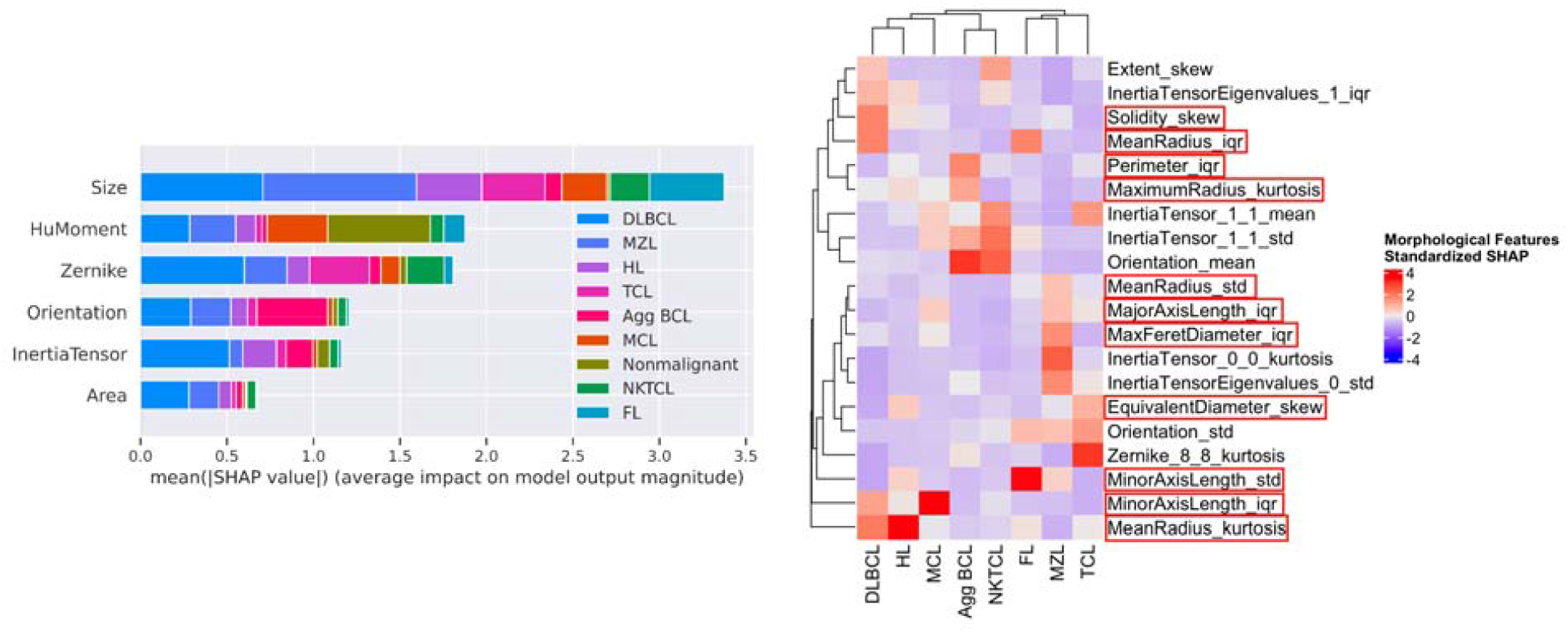
(SHAP): Feature importance analysis by SHapley Additive exPlanation (SHAP) values, while grouping related morphological features into different categories (left); the SHAP value breakdown of the top 20 morphological features in each diagnosis (right) with the most important features in the “size” group circled.

**Figure 5.**
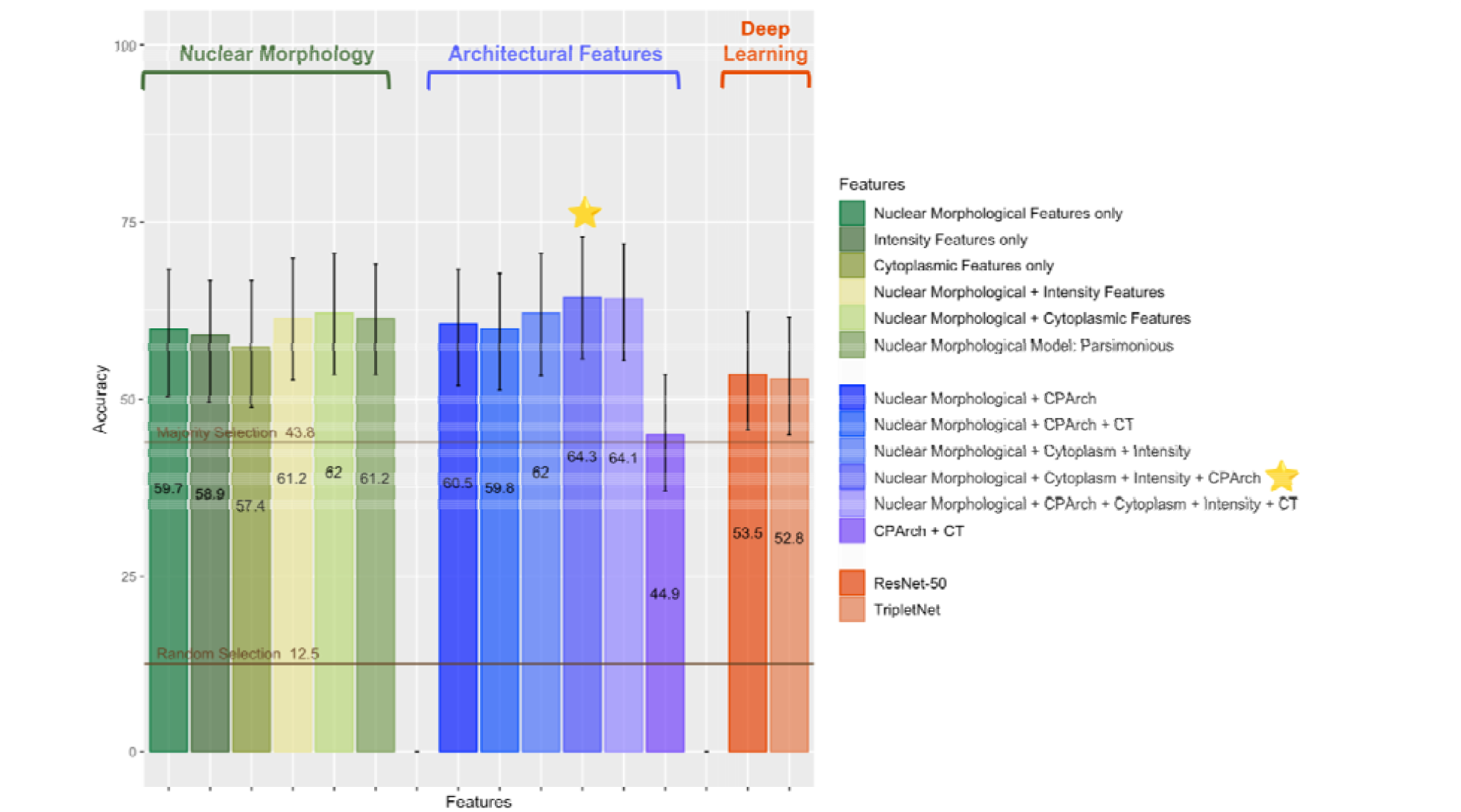
(Performance Summary): Performance summary for all models in Section 1 (Nuclear Morphology), Section 2 (Architectural Features), and Section 3 (Deep-Learning), with Majority Selection line representing a condition case when the model always predicts the most prevalent class and Random Selection line representing the model that performs random classification. The best performing model (referred to as “Best H&E Model” below) is marked with a star.

#### Section 1.2 Nuclear Architecture

We hypothesized that architectural characteristics such as clustering tendency among nuclei would improve accuracy on certain types of lymphoma over nuclear morphological features alone. Architectural features alone achieved 44.9% ([37.0%, 53.5%]) top-1 accuracy, providing no significant effect in predictions. The best model utilizing both architectural and nuclear features was “Nuclear Morphological + Cytoplasm + Intensity + CPArch’’ (referred to as “Best H&E Model” below). It achieved 64.3% ([55.8%, 72.9%]) top-1 accuracy, the highest accuracy among all models using H&E features, though statistical tests did not show evidence for it being significantly different from the Nuclear Morphological Model. We further examined its per-class performance to find that it achieved 71.0% ([50.0%, 87.0%]) F1 score in predicting MCL, 19.4% higher than the Nuclear Morphological Model’s performance (51.6% [27.6%, 71.0%]), though this difference did not achieve statistical significance.

#### Section 1.3 Comparison to Black-Box Features

We hypothesized that the interpretable machine learning models produced by LymphoML would outperform deep-learning methods (TripletNet and ResNet) used in prior studies given the scarcity of labeled examples. There was no significant difference between the test performance of the two deep-learning methods. TripletNet, which achieved better test accuracy than ResNet, was significantly inferior to the Best H&E Model produced using LymphoML. It was also worse than the Nuclear Morphological Model in both test accuracy and F1 score by a margin of ∼5% (top-1 test accuracy of TripletNet: 52.8% [44.9%, 61.4%], top-1 test accuracy of ResNet: 53.5% [45.7%, 62.2%]). For each class, the baseline generally had better performances.

### Section 2: Development of Best H&E-only Model and Performance Analysis

#### Section 2.1 Comparison with Pathologists and Hematopathologists

We compared the performance of the Best H&E Model from Section 1.2 with two pathologists on H&E-stained TMA cores and two pathologists on H&E-stained WSIs (Figure 6). Best H&E Model’s test accuracy (64.3% [55.8%, 72.9%]) surpassed performances of all pathologists and hematopathologists. Statistical tests verified that the best-performing H&E model achieved test performance non-inferior to the General Pathologist on WSIs and Hematopathologist 1 on TMAs, and there was no evidence of the Best H&E Model’s performance being statistically different from any pathologist’s performance (Supplementary Table 12). A similar conclusion can be derived from other metrics (sensitivity, specificity, and AUROC).

**Figure 6.**
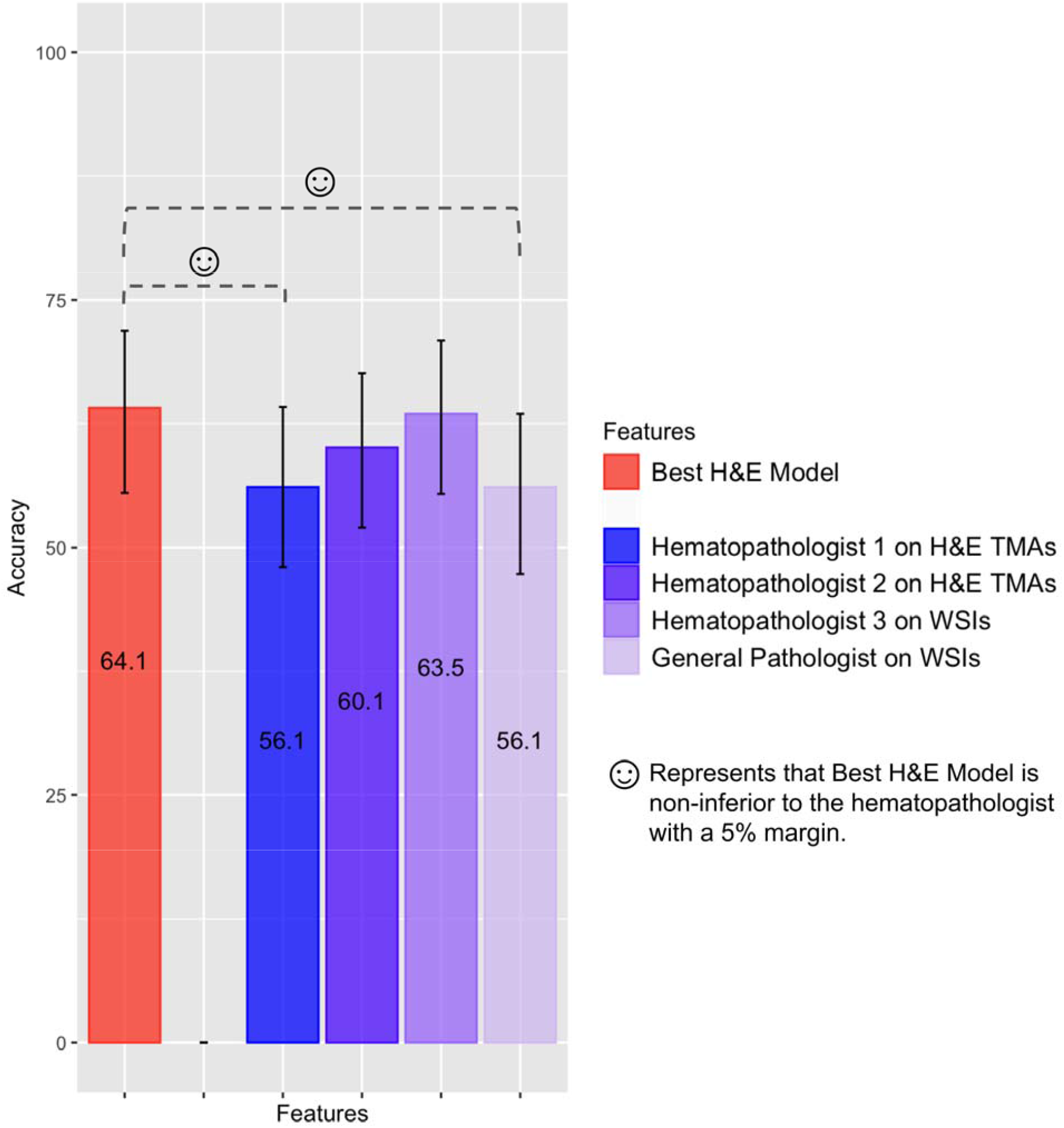
(Performance Comparison): Performance comparison between hematopathologists using TMAs, hematopathologists using WSIs, and the best-performing model using H&E TMAs that includes nuclear and cytoplasmic morphological, intensity, texture, and architectural features. Top-1 test accuracy and F1 scores are presented, with confidence intervals generated using 1000 bootstrapping samples from the test set.

We compared the per-class performance of our methods. On one hand, compared to pathologists’ predictions, the Best H&E Model failed to effectively identify any case in MZL and TCL while pathologists generally achieved ∼30% and 23.5% F1 scores in diagnosing MZL and TCL respectively. On the other hand, Best H&E Model achieved a 71.0% F1 score ([48.0%, 87.0%]) in MCL, surpassing all hematopathologists by a margin >18.8% and statistically superior to Hematopathologist 1 on H&E TMAs and Hematopathologist 3 on WSIs.

#### Section 2.2 Classification of Fewer Subtypes

In clinical practice, the distinction between DLBCL and other B-cell lymphomas frequently depends on morphologic features. Additionally, prior studies have shown excellent performance of machine learning tools in distinguishing DLBCL from non-DLBCL cases (Li et al. 2020; Miyoshi et al. 2020; Mohlman et al. 2020; Steinbuss et al. 2021; Achi et al. 2019). For this reason, we assessed performance of classification of DLBCL versus non-DLBCL in our cohort. As the diagnosis of DLBCL is defined by size (if the abnormal B-cells are at least the size of a histiocyte nucleus or at least two times the size of a normal lymphocyte), we initially considered models using only nuclear size/area features. Using only nuclear size/area features, the model achieved a test accuracy of 76.0% ([68.2%, 82.9%]) on DLBCL vs non-DLBCL; the most important nuclear features according to SHAP analysis were: maximum radius, mean radius, minor axis length, and nuclei area.

Our best model using nuclear morphologic, texture, intensity, and cytoplasmic features achieved a test accuracy of 79.8% ([72.9, 86.0]) on DLBCL vs non-DLBCL. We note that nuclear size features were among the most impactful features for this model too, e.g., the mean radius was the top feature according to SHAP analysis. Hematopathologists 1 and 2 achieved 73.0% and 73.0% test accuracy respectively using H&E TMAs only on DLBCL vs non-DLBCL. When using whole-slide images, Hematopathologist 3 achieved a top-1 test accuracy of 83.8%. By filtering patches whose maximum predicted score for any class was less than a specific threshold (i.e., the patch-based quality control (PQC) threshold used in (Steinbuss et al. 2021)), we boosted model performance to 87.0% on DLBCL vs non-DLBCL. However, note that in this approach, the model no longer outputs a prediction for TMA cores that did not contain at least one patch where the model’s maximum predicted score exceeded the threshold. Our selected threshold (0.9) resulted in the model only making a prediction for approximately two-thirds of the patients.

Next, we grouped lymphoma subtypes into categories that are similar in terms of clinical behavior and therapeutic approaches: B-cell lymphomas (DLBCL and Agg BCL), classic Hodgkin lymphoma, follicular and marginal zone lymphoma, mantle-cell lymphoma, and T-cell lymphomas (NKTCL and TCL). We re-trained our best model on the same dataset to classify patients into one of the aforementioned five groups. On this task, our best model achieved a test accuracy of 69.0% ([61.2, 77.5]), a five percent improvement over the model on the original eight-way classification task. In the fewer category setting, nuclei area shape features such as mean radius and minor axis length remained the top-performing features according to SHAP analysis.

#### Section 2.3 Additional Stains

The ground truth diagnosis for our cohort was based on review by hematopathologists of the H&E-stained whole-slide images and a panel of 46 immunohistochemical (IHC) stains that were performed on all cases. The panel of IHC stains was chosen for both diagnostic and research purposes and thus for purely diagnostic purposes, this set of IHC stains is more extensive than what is necessary for diagnosis in the vast majority of cases. For each case and each immunostain, a hematopathologist scored the lymphoma cells as positive or negative for the immunostain (e.g. CD30-positive) or “cannot interpret.” As the hematopathologist-scored IHC results were available for our study, we set out to determine the diagnostic accuracy of using this IHC information alone, the diagnostic accuracy of using a limited set of IHC stain results, and the diagnostic accuracy of using a limited set of IHC stain results in conjunction with the H&E-based model.

Out of the 46 different immunostains available in our dataset, we considered six candidate immunostains based on the authors’ impression of markers that would be the highest yield in arriving at a correct diagnosis considering the diagnostic categories in our cohort: CD10, CD20, CD3, EBV-ISH, BCL1/cyclin D1, and CD30. The diagnostic power of the selected stains was also verified experimentally by checking that most of them rank top 20 among all stains in importance (see Figure 8). For each candidate immunostain, the pathologist’s score was included as an additional categorical feature to the Best H&E Model. We also grouped lymphoma subtypes into the five clinically similar categories and evaluated the accuracy and F1 scores of models using features extracted from H&E stains along with different combinations of immunostain indicator features. The baseline model using 46 different immunostains (and no H&E) achieved a top-1 accuracy of 86.1% ([79.8%, 91.5%]). Using the six selected immunostains alone, without the H&E, the model achieved a top-1 accuracy of 75.2% ([67.4%, 82.2%]), which is statistically inferior to the model using all 46 immunostains (see Figure 7). The Best H&E Model (Nuclear Morphological + Intensity + Cytoplasm + CPArch) augmented by six selected immunostains (CD10, CD20, CD3, EBV ISH, BCL1, and CD30) achieved a top-1 accuracy of 85.3% ([79.1%, 90.7%]) and showed no evidence of difference from the model using all 46 immunostains.

**Figure 7.**
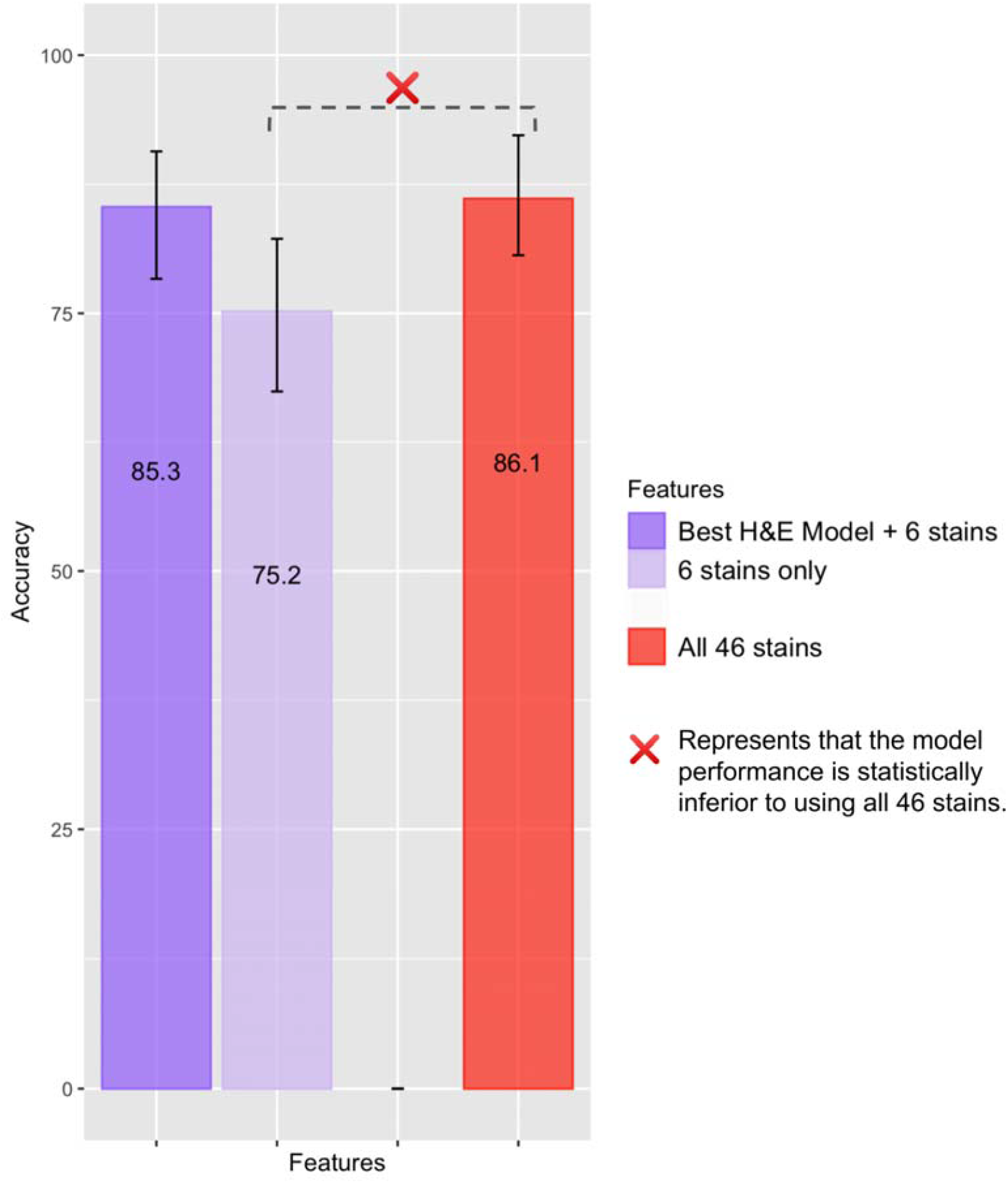
(Immunostains): Performance comparisons of models using features extracted from H&E and 6 selected immunostains to the baseline consisting of all 46 stains.

**Figure 8.**
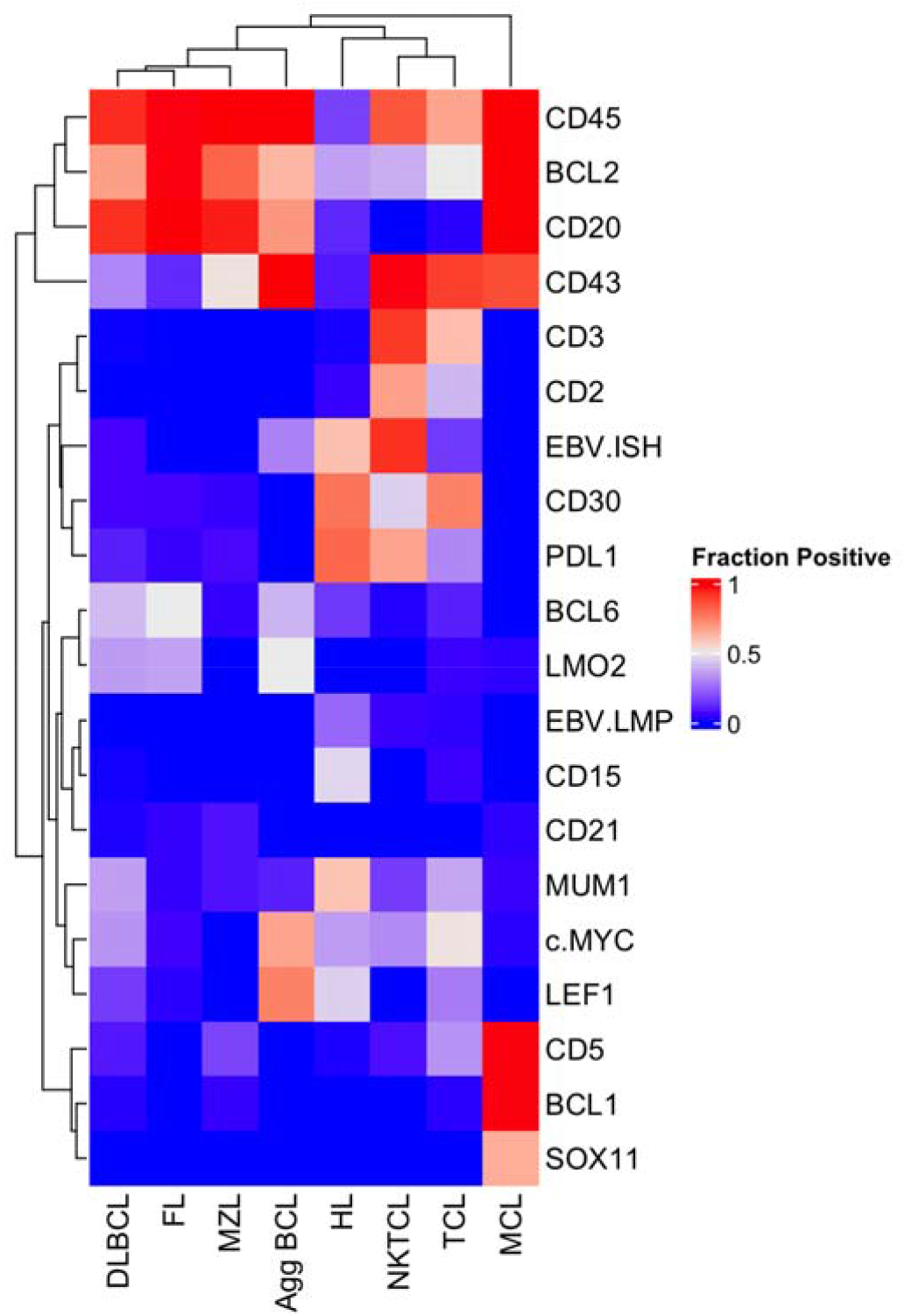
(Immunostains Importance): The 20 Immunostains with the highest SHAP values and their diagnostic power in clinical practice (illustrated by fraction of positive stains).

## Discussion

In this study, we assessed the performance of interpretable and deep-learning approaches in the classification of eight lymphoma categories using a cohort of 670 lymphoma cases. To date, this cohort represents the second-largest group of lymphomas studied using machine learning methods and this study represents the most comprehensive interpretable morphometric analysis of lymphomas. Our main finding is that, using only H&E-stained TMA material, LymphoML achieves performance comparable to that of experienced hematopathologists reviewing only the H&E-stained tissue. The highest performance in the diagnostic classification task was achieved using an interpretable model. We find that combining information from the H&E-based model together with the results of a limited set of IHC stains resulted in a similar diagnostic accuracy as with a much larger set of IHC stains.

Prior studies that applied machine learning methods to lymphoma diagnosis have achieved accuracies of 94% to 100% (Janowczyk and Madabhushi 2016; Brancati et al. undefined 2019; Steinbuss et al. 2021; Zhang et al. 2020; Li et al. 2020; Miyoshi et al. 2020; Mohlman et al. 2020; Achi et al. 2019). Although they have demonstrated high accuracy, it is not clear how these tools can be employed in the real-world given the limited number of diagnostic categories studied. Supplementary Table 5 summarizes notable features from these prior studies. We note that a recently presented abstract used 4247 H&E-stained WSIs of 8 lymphoma types to train and test a neural network, achieving an overall diagnostic accuracy of 81% (Zhu et al. 2022). Though the diagnostic accuracy of machine learning algorithms may decrease as the number of diagnostic categories increases, workflow models that leverage the power of machine learning to maximize the yield of the H&E-stained slide may yet prove useful if they can reduce the number of IHC stains necessary to arrive at a diagnosis.

Our study differs from prior published studies in several notable ways. Firstly, our study was performed on a cohort that contains 2- to 4-fold more diagnostic categories than all prior published studies. Though our cohort contains the second-largest number of cases thus far used to develop machine learning tools for lymphoma diagnosis, the number of cases is within the same order of magnitude as that used in other studies. In real-world settings with a large number of possible diagnoses, it is likely that deep-learning methods will require a proportionate increase in the number of cases to perform well. With a limited number of examples for specific diagnostic categories, models can only learn a small range of the potentially wide morphological variability present in these subtypes. Therefore, feature-engineering approaches may yet provide superior diagnostic yield when the number of cases per diagnosis is insufficient to use deep-learning. Additionally, though the TMA cores are highly enriched for lymphoma, they contain variable amounts of background cells and extracellular tissue (e.g. collagen). In our study, we did not specifically annotate lymphoma within each core and thus the regions of interest used for our algorithms are not as enriched as in the study published by Steinbuss et al, who used TMAs and had pathologists annotate the region with lymphoma within the TMA core. It is notable that our interpretable models, developed on a limited volume of tissue, achieved the same diagnostic accuracy as hematopathologists using whole-slide images. Our study suggests that computational tools may be able to extract more diagnostic information from less tissue than a pathologist. The growing use of needle core biopsies in clinical practice makes judicious use of tissue even more critical. Computational tools that maximize the diagnostic yield of the H&E-stained slide could potentially reduce the number of ancillary stains and thus avoid the need for repeat biopsy or an excisional biopsy.

Pathologists use features similar to the nuclear shape features identified by SHAP including nuclear-to-cytoplasmic ratio, nuclear contour irregularities, and nuclear size as one of many clues to determine if cells are normal or malignant. DLBCL, by definition, consists of sheets of large B-cells, with ‘large’ defined as nuclei that are at least the size of a histiocyte nucleus or two-times the size of a normal lymphocyte. Consistent with this definition used to render the ground truth diagnosis, our interpretable model showed that nuclear size features differed between DLBCL and other lymphoma subtypes. Specific features that provided the greatest diagnostic yield for 8-way classification included mean radius, minor axis length, maximum Feret diameter, solidity, and orientation. In the two-way classification of DLBCL versus non-DLBCL, the most important nuclear features were mean radius, maximum radius, and variation in bounding box size. The fact that our model identified nuclear features as most important in distinguishing DLBCL from non-DLBCL is entirely consistent with the World Health Organization definition of DLBCL. Our work represents the first pixel-level assessment of the size and shape differences between DLBCL and non-DLBCL. Prior studies that quantitatively assessed interpretable morphologic features in lymphoma diagnosis have used a smaller number of cases and fewer diagnostic categories (Gupta et al. 2010; Lesty et al. 1986; W.-H. Yu et al. 2021).

Based on our statistical test results, there is no significant improvement in prediction performance after adding architectural features relative to the Nuclear Morphological Model. Compared with carcinomas that form higher-order structures such as tubules, the higher-order spatial relationship between cells in lymphomas may be less diagnostically informative. Notable architectural features that can be helpful in lymphoma diagnosis include a nodular architecture (as seen in FL, MCL, MZL) and proliferation centers in CLL/SLL. In CHL, large atypical cells are surrounded by smaller lymphocytes, eosinophils, and neutrophils and thus the spatial relationship between the cells in the infiltrate represents an important clue to the diagnosis. There are several reasons why spatial features may not provide significant diagnostic yield in our study even though pathologists find spatial features to be of significant diagnostic relevance. The higher-order architectural features that are helpful to pathologists (e.g. nodularity, proliferation centers) are not as well represented in TMA cores as they are in whole tissue sections. Also, tumor cells and the immune microenvironment in lymphomas are derived from the same cell types, making their distinction from one another difficult. In the successful example of using architectural information from Ripley’s K statistic in Subramanian et al., authors manually designed thresholds based on cell areas and cell average intensity to categorize cells into two groups and computed cross-Ripley’s K statistics. This is a non-trivial step for lymphoma diagnosis. By categorizing cells into “large” and “small” based on cell areas in immunostains experiments, cross-Ripley’s K statistics show no effect in improving model prediction. To fully utilize the potential of architectural information in statistics like Ripley’s K, more carefully designed thresholds are necessary and this can become increasingly labor-consuming with more classes and subtle differences between classes. We conclude that the main power in driving lymphoma prediction accuracy using H&E-based models still comes from nuclear morphological features.

In order to classify lymphoma subtypes, information within tumor regions is discriminatory. Hence when using deep-learning methods to classify WSIs into tumor subtypes, MIL and methods like CLAM use fixed or learned aggregation schemes of patch-level predictions to focus on regions of interest (which generally correlated with tumor regions in classification tasks). In other cancer indications, this is performed more explicitly by training a tumor segmentation model on the WSI and only using patches in the tumor to train a deep-learning model. Coudray et. al. trained a model on tumor patches to predict gene mutation in lung adenocarcinoma and lung squamous cell carcinoma (Coudray et al. 2018). Similar methods have been used in ovarian carcinoma (K.-H. Yu et al. 2020), lymphoma (Achi et al. 2019), and hepatocellular carcinoma. Since TMAs contain tissue highly enriched for tumor and are extracted from regions in the WSI with high tumor burden, our method is, in principle, equivalent to applying tumor segmentation and only extracting tumor-rich patches from the WSI. However, since the TMA is a small subset of the tumor regions of a WSI, we have a much smaller tumor volume per sample. This results in a smaller number of patches than other deep-learning studies with WSIs. Our cohort only contains 670 cases distributed across eight lymphoma subtypes, while previous studies “on lymphoma subtyping included between 34 and 259 cases per entity and a total of 2560 to 850,000 image patches” (Steinbuss et al. 2021). Due to the heavy label imbalance, we only extracted over 10,000 patches for two subtypes: DLBCL (32,926) and CHL (11,829) (Supplementary Table 8), which were the best-performing categories. Other works using WSIs such as (Chen et al. 2021) for adenocarcinoma and squamous cell carcinoma classification had an order of magnitude more data: 9662 WSIs with number of pixels at least 1000 times greater than that of ImageNet images. The authors found that reductions in training dataset size and image magnification level reduced model performance on the test set. This strongly suggests that deep-learning models underperformed due to an insufficient number of patches for labels other than DLBCL and CHL.

Next, we discuss why adopting patch-level model training and merging patch-level predictions via plurality vote is appropriate for our study. As TMA cores are highly enriched for lymphoma and are extracted from regions in the WSI with high tumor burden, the vast majority of tissue in each core is correlated with its corresponding lymphoma subtype and multiple instance learning is not strictly necessary. However, a small number of tumor cells per patch could be a limiting factor, and the minimal number of tumor cells per patch needed for a reliable result is currently not clear (Steinbuss et al. 2021). There is no standard image patch size (Steinbuss et al. 2021) so we performed patch-resolution experiments to select the best patch size for feature-based models (at the extreme, using one patch to represent the entire core). Results are reported in Supplementary Table 14. We found experimentally that extracting a small number of patches per core (specifically, 4 patches per core) led to the best model performance, and in particular, better performance than core-level model training and prediction.

The cohort used in this study contained immunophenotypic data that had been used to arrive at the ground truth diagnosis. This immunophenotypic data was based on IHC stains performed on TMA sections reviewed by a hematopathologist, who determined if the lymphoma cells were positive or negative for the marker. These data allowed us to compare the H&E-based model with models only based on the IHC score result. Using data for all 46 IHC stains provided a baseline for the diagnostic accuracy of using only IHC-stains as interpreted by a pathologist. We next measured the diagnostic accuracy of using a limited set of IHC stains and assessed the impact of adding the H&E-based model to the limited IHC set. Adding the H&E-based model to six selected IHC stains achieved a diagnostic accuracy not statistically different from a model using all 46 IHC stains.

We note that 46 immunostains represent a marked excess of stains compared with what is typically necessary to arrive at a diagnosis. In general, fewer than 10 immunohistochemical stains are necessary to make a diagnosis of lymphoma. The diagnostic accuracy of using 46 immunostains represents an artificial scenario that provides a baseline comparison condition, not a comparison with routine clinical practice. It answers the question: How accurate is a model that simply incorporates immunophenotype? It is known that this model will be imperfect as the definition of DLBCL is based on cell size as assessed on an H&E-stained section. A more complicated question is whether features identified on H&E-stained sections are sufficient to reduce the number of immunohistochemical stains necessary to arrive at a diagnosis. In our study, we provide the first demonstration that a model that incorporates immunophenotypic features and features extracted from H&E-stained sections can achieve the same diagnostic accuracy as a model that uses a larger number of immunostains. Though machine learning tools will no doubt maximize the diagnostic yield from H&E-stained slides, there will likely be many situations in which a definitive diagnosis cannot be made by only using the information present on the slides. Models that combine machine learning assessment of H&E-stained tissue together with immunophenotypic information merit further assessment as a way to incorporate machine learning tools into diagnostic practice.

As H&E-stained slides are cheaper than IHC stains by at least an order of magnitude, extracting the maximal diagnostic yield from H&E-stained slides can reduce the number of IHC stains ordered and thus reduce costs without a reduction in diagnostic accuracy. Additionally, we note that pathologists usually order a custom panel of IHC stains for each case. In our study, we showed that models combining H&E features with a *standard* set of six IHC stains can arrive at the correct diagnosis with a high degree of confidence in the majority of cases (85.3%). Standardizing the panel of IHCs reduces variability in practice and improves cost effectiveness. This portion of our study suggests a novel way to incorporate machine learning tools into fields of pathology that require immunophenotyping for definitive diagnosis. Often, more experienced or subspecialty pathologists more efficiently arrive at a diagnosis than a general pathologist, but in many fields of pathology, immunophenotyping (with immunohistochemistry or flow cytometry) is critical for diagnosis, prognosis, and therapy selection. If a less-experienced or general pathologist encounters a case, machine learning tools that maximize the diagnostic yield from the H&E-stained slide may reduce the number of ancillary studies necessary to achieve the same level of diagnostic accuracy. As hardware and software are likely easier to scale than immunohistochemistry and flow cytometry, this approach to deploying machine learning tools in the field of pathology may reduce the overall cost of pathology. Just as more experienced pathologists arrive at a correct diagnosis with fewer immunohistochemical stains because they obtain more diagnostic yield from the H&E image, computational tools that extract the maximum diagnostic yield from H&E images may reduce the number of immunohistochemical stains necessary to achieve the same diagnostic accuracy.

### Strengths

Our study has several strengths. First we found that the best models derived using LymphoML method achieved top-1 test accuracy equivalent to that of experienced hematopathologists even when classifying among many more (eight) lymphoma subtypes than other studies. Using feature importance analysis, our computational methods can help pathologists better understand the primary characteristics of the lesion that contribute to the model’s prediction. Second, our study cohort was obtained from patients in Guatemala, a patient population that is not represented in current datasets in the field of digital pathology. Thus, the cases generated from our study will likely prove valuable in the effort to build diverse datasets for further development of computational tools in the field of medicine. Most prior works exploring computational tools for classifying lymphomas focused on the use of whole-slide images, often with expensive, manually-generated patch-level annotations. WSIs often require manual annotations because these larger images usually contain both cancerous and normal surrounding tissue. Here, we use TMAs, a strength and limitation as other studies (Beck et al. 2011) have discussed. TMAs tend not to require expensive manual annotations because cores are already enriched for lymphoma. Using TMAs, computational tools are likely more cost-effective and usable in low-middle income countries where acquisition of labeled data can prove costly.

### Limitations

LymphoML has several limitations that will need to be addressed before the resulting models can be applied in clinical practice. First, the data were collected and processed in a single institution by a single slide scanner. Staining TMAs for H&E and the immunostains were also performed at the same location. We should evaluate the model’s generalizability on cohorts from other institutions collected using different technical setups and/or slides scanned on different machines. Next, TMAs capture only a small portion of the full tumor volume and are much smaller than WSIs (Beck et al. 2011). Thus, we could have likely trained more powerful models by analyzing WSIs. We should also incorporate benign cases to develop models that can distinguish lymphoma from non-lymphoma, another critical real-world distinction to supplement pathology review of H&E. The heavy class imbalance meant that only a small number of examples were available for Agg BCL, MZL, TCL (<10 patients). A limited number of patches per core can only display a small subset of the wide morphological variability present in Agg BCL, MZL, and TCL, so it cannot be expected that our algorithms will reliably classify these subtypes. Finally, to implement LymphoML in a clinical setting, pathologists would have to suspect one of the validated diagnostic categories, select a region of interest, and then the model would render a favored diagnosis. Alternatively, future studies with larger data sets, more diagnostic categories, and a more diverse set of background tissues may someday enable automated identification of target lesion(s) and subsequent diagnostic categorization.

In summary, we proposed LymphoML, an interpretable machine learning approach that can classify a wide range of lymphoma subtypes using H&E-stained tissue microarray material and evaluated the features that provide the highest diagnostic yield. Our study highlights the potential of interpretable machine learning toolkits in lymphoma subtype classification and in the diagnosis of other cancer types, as well as the general promise of AI-assisted workflows in diagnostic pathology.

## Data Availability

The datasets used and/or analyzed during the current study are available from Sebastian Fernandez-Pol on reasonable request. We are acquiring a Data Use Agreement (DUA) to be able to share the raw data.

## Code Availability

The code developed during the study can be downloaded from the following Github link: https://github.com/rajpurkarlab/LymphoML.

## Data Availability

All data produced in the present study are available upon reasonable request to the authors.

## Author Contributions

X.Y., V.S., V.K., S.F., and P.R. developed the concept and design; X.Y., V.S., V.K., S.F., P.R., R.R., O.S., R.R., F.V., F.V., E.L.B., D.M.W., and Y.M. performed acquisition, analysis, or interpretation of data; A.Y.N., S.F., and P.R. provided supervision. X.Y., V.S., V.K., S.F., and P.R. drafted the manuscript, and all authors provided critical revision of manuscript for important intellectual content.

## Ethics Declaration

### Ethics Approval

This study was approved by the institutional review board of Stanford University.

